# Potential for bias in (sero)prevalence estimates when not accounting for test sensitivity and specificity

**DOI:** 10.1101/2022.11.24.22282720

**Authors:** Sarah R Haile, David Kronthaler

## Abstract

**Background:** The COVID-19 pandemic has led to many studies of seroprevalence. A number of methods exist in the statistical literature to correctly estimate disease prevalence or seroprevalence in the presence of diagnostic test misclassification, but these methods seem to be less known and not routinely used in the public health literature. We aimed to examine how widespread the problem is in recent publications, and to quantify the magnitude of bias introduced when correct methods are not used.

**Methods:** A systematic review was performed to estimate how often public health researchers accounted for diagnostic test performance in estimates of seroprevalence. Using straightforward calculations, we estimated the amount of bias introduced when reporting the proportion of positive test results instead of using sensitivity and specificity to estimate disease prevalence.

**Results:** Of the seroprevalence studies sampled, 78% (95% CI 72% to 82%) failed to account for sensitivity and specificity. Expected bias is often more than is desired in practice, ranging from 1% to 12%.

**Conclusions:** Researchers conducting studies of prevalence should correctly account for test sensitivity and specificity in their statistical analysis.

## Introduction

Since the beginning of the SARS-CoV-2 pandemic, thousands of papers have been published detailing seroprevalence estimates in various populations (1). A glance into recent publications indicates that while some researchers used simple approaches such as proportions or logistic regression, others used complicated methods like Bayesian hierarchical models. An important question is therefore how often these methods are used in epidemiological studies and what, if any, degree of bias was introduced by using one method or the other.

As diagnostic tests are not 100% accurate, it is expected that some small number of test results will be either false positives or false negatives. Using a simple proportion of the number of positive diagnostic tests over the total number of tests ignores any misclassification inherent to the test. In the case where there are similar numbers of true positives and true negatives in the population, the bias introduced by using the proportion of positive tests to estimate the proportion of subjects with the disease may not be very high. However, if the rate of false positives differs greatly from that of false negatives, the bias may be quite large.

For example, in Table 1, 35.5% (355/1000) of subjects had a positive test result, but the true disease prevalence in this population is 30.0% (196/1000). So there is a bias of 5.5% because there are many more false positive test results (n = 70) than false negatives (n = 15). Other examples of this phenomenon are found in the literature (2,3). Statisticians often talk about sensitivity, 95% in this example, and specificity, 90%, of the diagnostic in relation to these quantities (described in more detail below), but it is accepted that without a “gold standard” diagnostic tool, it is difficult to accurately assess disease prevalence.

**Table 1:** Typical example of 2×2 table comparing diagnostic test results and disease status.

|  | Test neg | Test pos | total |
| --- | --- | --- | --- |
| Disease neg | 724 | 80 | 804 |
| Disease pos | 20 | 176 | 196 |
| total | 744 | 256 | 1000 |

Accounting for such misclassification in the interpretation of diagnostic tests is certainly not new in the literature. A straightforward method of adjusting observed prevalence is available (4,5), which gives a maximum likelihood estimate of true prevalence assuming predefined test sensitivity and specificity. The Rogan-Gladen correction has been extended to compute confidence intervals (6,7). Recently, an adaptation of the Rogan-Gladen correction that accounts for sampling bias, for example if only hospitalized subjects as opposed to the general population have been tested, has been proposed (8–10). Bayesian approaches have also been developed (3,11,12). A comparison of Bayesian and frequentist methods (13) showed that Bayesian methods are to be preferred, or the method of (4) with confidence intervals of (7).

Despite this extensive treatment of the misclassification problem in the statistical literature, many public health researchers appear to not realize they may be publishing biased results or know what to do about it. In what follows, a systematic review quantifies the proportion of recent publications estimating seroprevalence that do not correct for diagnostic test performance. We will describe key concepts, and derive an estimate of the bias, as well as a range of prevalences where such naive estimates show low bias. Bias estimates will be described according to test sensitivity and specificity, and we will apply these results to a real example of SARS-CoV-2 seroprevalence in children.

## Methods

To start, we introduce some notation. Disease status, *D*, is denoted 1 if a subject has the disease in question (or for the case of seroprevalence, has antibodies for it), and 0 otherwise. Similarly, the result of the diagnostic test, *Y*, is given as 1 if the subject tests positive for the disease, and 0 otherwise. FP is often used to refer to false positive test results, and similarly FN for false negatives, TN for true negatives and TP for true positives.

Prevalence is the probability of having the disease of interest, *P* = *Pr*(*D* = 1). Often in prevalence studies, this probability is studied at a specific point in time, giving so-called point prevalence (14). Seroprevalence, a related concept, looks at the proportion of individuals in the population have antibodies for a specific disease, for example, SARS-CoV-2 (15). Sensitivity, denoted *Se*, sometimes also called the true positive fraction (TPF), is the probability of having a positive test result, given that the subject has the disease, *Pr*(*Y* = 1|*D* = 1) (2). On the other hand, specificity, *Sp* is the probability of having a negative test result when a subject does not have the disease, *Pr*(*Y* = 0|*D* = 0) (sometimes 1 – specificity is discussed, which is often referred to as false positive fraction, or FPF (16)). In real settings where true disease status is known via another method, sometimes referred to as the “gold standard”, *Se* can be computed as *TP*/(*TP* + *FN*), where TP is the number of true positives and FN is the number of true negatives. Similarly, *Sp* can be computed as 1 − *FP*/(*FP* + *TN*).

The proportion of positive tests can be expressed as

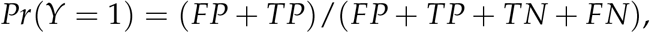

while the disease prevalence in the sample can be expressed as

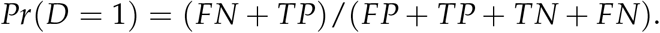

The difference between these two quantities is simply (*FP* − *FN*)/(*FP* + *TP* + *TN* + *FN*), that is, the proportion of false positives minus the proportion of false negatives.

According to the definition of joint probability *Pr*(*A, B*) = *Pr*(*A*|*B*)*Pr*(*B*), the proportion of false positives can be written as

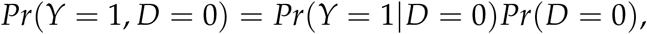

which simplifies to (1 − *P*)(1 − *Sp*). In a similar fashion, the proportion of false negatives can be written as

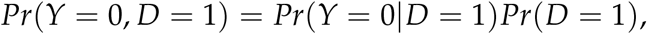

which simplifies to *P*(1 − *Se*). The bias when using the proportion of positive tests, *Pr*(*Y* = 1), to estimate the proportion with disease, *Pr*(*D* = 1), is therefore (1 − *P*)(1 − *Sp*) − *P*(1 − *Se*) or equivalently 1 − *Sp* + *P*(*Sp* + *Se* − 2).

Suppose we want to guarantee that the bias is no larger than, say, *δ* = 0.02, that is ±2% in either direction. We can solve

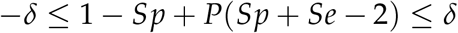

for *P*, to get:

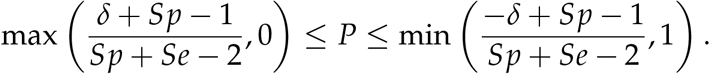

The lower bound will be 0 if *δ* ≥ 1 − *Sp*, while the upper bound will be 1 if *δ* ≥ 1 − *Se*. Therefore, if both *Se* and *Sp* are very high, say 99% or higher, then the proportion of positive tests is a good estimate of the true prevalence. If only *Se* is that high, this is will be true only when the true prevalence is quite high, and conversely if only *Sp* is very high, this will be true only when true prevalence is quite low. When neither *Se* nor *Sp* is high, the proportion of positive tests may or may not be a good estimate of the true prevalence.

One simple way to reduce this bias, if no dependence on covariates is assumed, is to use the Rogan-Gladen correction (4). Assuming an observed fraction *P*_*obs*_ of positive test results, the corrected prevalence is

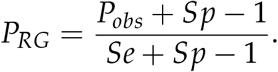

In a small number of cases, primarily when the sample size and the prevalence are both small (17,18), the Rogan-Gladen correction will yield values less than 0 or greater then 1. However, even if this “clipped” version has some bias, the variance will be smaller.

The systematic review of recent studies of seroprevalence in the literature started with a pubmed (https://pubmed.ncbi.nlm.nih.gov/) search for “covid-19 seroprevalence”, which yielded 637 publications published in 2022. Publications were included in the systematic review if they assess COVID-19 seroprevalence in humans, and were published in 2022 in English or German. Exclusion criteria included: 1) studies comparing seroprevalence in different subgroups, 2) studies examining risk factors for seropositivity, 3) studies in animals, 4) reviews, 5) methodological papers, 6) studies with possible conflict of interest, 7) if the full text was not available or 8) if the publication was a research letter. The following information was extracted: 1) whether the aim of the study was to assess COVID-19 seroprevalence in humans, 2) the sensitivity and 3) specificity of the diagnostic test, 4) the reported seroprevalence estimate (the first mentioned value, and if unadjusted was reported before adjusted, we extracted the most adjusted value of the first mentioned seroprevalence), and 5) which statistical methods were used to calculate seroprevalence. A protocol for the systematic review was developed using the PRISMA-P checklist (https://osf.io/b59x2/). Two independent reviewers (SRH and DK) screened the publications using the `rayyan.ai` web-based tool, and performed data extraction in parallel using a structured spreadsheet. Discrepancies were resolved by discussion. Summary statistics were computed for the methods used (n (%)), reported sensitivity and specificity (median [range]) and estimated bias (median [range]).

To provide a concrete example of this problem, we use the Ciao Corona study (19), a school-based longitudinal study of seroprevalence in Swiss school children with 5 rounds of SARS-CoV-2 antibody testing between June 2020 and June 2022, covering a range of seroprevalences in the population (Trial Registration: ClinicalTrials.gov NCT04448717). The study was conducted in accordance with the Declaration of Helsinki and approved by the Ethics Committee of the Canton of Zurich, Switzerland (2020-01336). All participants provided written informed consent before being enrolled in the study.

## Results

To examine the methods actually used in seroprevalence studies in the literature, we performed a systematic review of publications from 2022 which estimated COVID-19 sero-prevalence in humans (Table 2). Of the 640 publications identified in pubmed, 4 were duplicates, and 349 were excluded (5 represented possible conflicts of interest, 7 were published in languages other than English or German, 2 did not examine COVID-19, 9 were animal studies, 22 described secondary research, 233 did not assess seroprevalence, 41 compared subgroups or risk factors for seropositivity, 3 did not have full texts available, and 23 were published as research letters). Of the remaining 291 publications (Supplementary Material Table S1), 77.7% (n = 226, 95% CI 72.4% to 82.3%) did not adjust for diagnostic test performance, while 22.3% corrected for sensitivity and specificity of the diagnostic test (n = 65, 17.7% to 27.6%). Among the publications which adjusted for test characteristics, 39 (13.0%) used Rogan-Gladen correction, 18 (6.2%) used Bayesian approaches, and 8 (12%) mentioned adjustment but did not specify further.

**Table 2:**
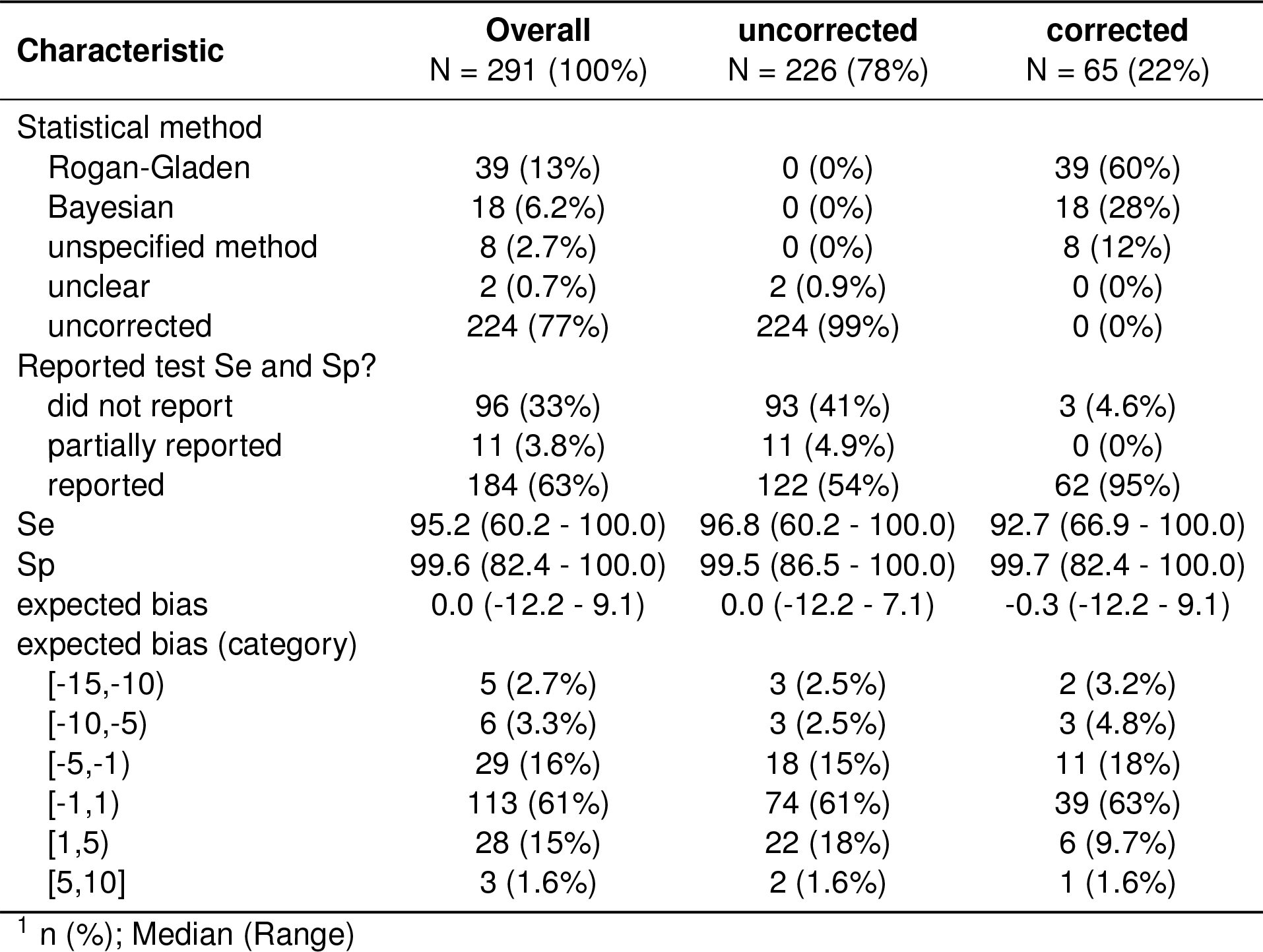
Key outcomes of systematic review. The main analysis included 291 publications meeting all inclusion criteria.

| **Characteristic** | **Overall**<br>N = 291 (100%) | **uncorrected**<br>N = 226 (78%) | **corrected**<br>N = 65 (22%) |
| --- | --- | --- | --- |
| Statistical method |  |  |  |
| Rogan-Gladen | 39 (13%) | 0 (0%) | 39 (60%) |
| Bayesian | 18 (6.2%) | 0 (0%) | 18 (28%) |
| unspecified method | 8 (2.7%) | 0 (0%) | 8 (12%) |
| unclear | 2 (0.7%) | 2 (0.9%) | 0 (0%) |
| uncorrected | 224 (77%) | 224 (99%) | 0 (0%) |
| Reported test Se and Sp? |  |  |  |
| did not report | 96 (33%) | 93 (41%) | 3 (4.6%) |
| partially reported | 11 (3.8%) | 11 (4.9%) | 0 (0%) |
| reported | 184 (63%) | 122 (54%) | 62 (95%) |
| Se | 95.2 (60.2 - 100.0) | 96.8 (60.2 - 100.0) | 92.7 (66.9 - 100.0) |
| Sp | 99.6 (82.4 - 100.0) | 99.5 (86.5 - 100.0) | 99.7 (82.4 - 100.0) |
| expected bias | 0.0 (-12.2 - 9.1) | 0.0 (-12.2 - 7.1) | -0.3 (-12.2 - 9.1) |
| expected bias (category) |  |  |  |
| [-15,-10) | 5 (2.7%) | 3 (2.5%) | 2 (3.2%) |
| [-10,-5) | 6 (3.3%) | 3 (2.5%) | 3 (4.8%) |
| [-5,-1) | 29 (16%) | 18 (15%) | 11 (18%) |
| [-1,1) | 113 (61%) | 74 (61%) | 39 (63%) |
| [1,5) | 28 (15%) | 22 (18%) | 6 (9.7%) |
| [5,10] | 3 (1.6%) | 2 (1.6%) | 1 (1.6%) |

<sup>1</sup> n (%); Median (Range)

**Table 3:** Estimated bias (in percentage points) for selected combinations of sensitivity (Se), specificity (Sp) and disease prevalence (P)

| Se | Sp | P=2 | P=10 | P=30 | P=50 | P=90 | P=98 |
| --- | --- | --- | --- | --- | --- | --- | --- |
| 90 | 99 | 0.8% | -0.1% | -2.2% | -4.4% | -8.8% | -9.8% |
| 90 | 95 | 4.7% | 3.5% | 0.5% | -2.2% | -8.5% | -9.7% |
| 90 | 90 | 9.8% | 8.0% | 3.8% | 0.1% | -7.6% | -9.6% |
| 95 | 99 | 0.9% | 0.4% | -0.8% | -2.1% | -4.4% | -4.9% |
| 95 | 95 | 4.8% | 4.0% | 1.8% | 0.0% | -4.0% | -4.8% |
| 95 | 90 | 9.7% | 8.7% | 5.2% | 2.7% | -3.6% | -4.7% |
| 99 | 99 | 1.0% | 0.8% | 0.4% | 0.0% | -0.8% | -1.0% |
| 99 | 95 | 4.9% | 4.4% | 3.3% | 1.9% | -0.3% | -0.9% |
| 99 | 90 | 9.8% | 9.0% | 6.8% | 4.6% | 0.1% | -0.8% |

Further, among those publications that did adjust for test performance, 122/226 (54.0%) reported sensitivity and specificity, the remaining publications either did not report test characteristics (41.0%, n = 93) or only reported partial test characteristics (4.9%, n = 11). Among all publications reviewed, it is therefore observed that 33% (99/291) neither adjusted for test performance nor reported sensitivity and specificity. Among those that did not correct for test performance but did report both sensitivity and specificity (n = 122), expected bias ranged from -12.2% to 7.1%. 74 (61%) of the publications reporting seroprevalence to within ±1% of the true value despite not using any adjustment, while the remaining 48 (39%) needed adjustment for test performance (8 of those were not even within ±5%). It could be inferred therefore that approximately 41 of the 104 publications not or partially reporting test performance are also in need of adjusted seroprevalence estimates to account for test performance, even though all of those publications reported naive estimates. These results did not change when including publications denoted “research letters” (Supplementary Material Table S2). While the need to adjust seroprevalence estimates for test performance is well known the the statistical literature, the vast majority of published analyses on this topic fail to account for it when they should have. This problem is also not restricted to “low quality” journals, as such analyses can be found also in many prominent journals (Supplementary Data).

Next, we sought to characterize scenarios where expected bias would be minimal. Using the result bias = 1 − *Sp* + *P*(*Sp* + *Se* − 2) described above, we calculated the expected bias for a range of reasonable combinations of sensitivity, specificity and disease prevalence (Table 2, Figure 1). When sensitivity and specificity were both 90%, bias was as high as 10%, especially near prevalences of 0% or 100% (bottom row of Table 2, solid line in leftmost panel of Figure 1). When specificity was 90%, a bias of 10% could be expected with small prevalences near 0% even if sensitivity was 99% (e.g. 3rd line of Table 2). The least bias, 1%, could be expected where sensitivity and specificity were both 99% (1st line of Table 2).

**Figure 1:**
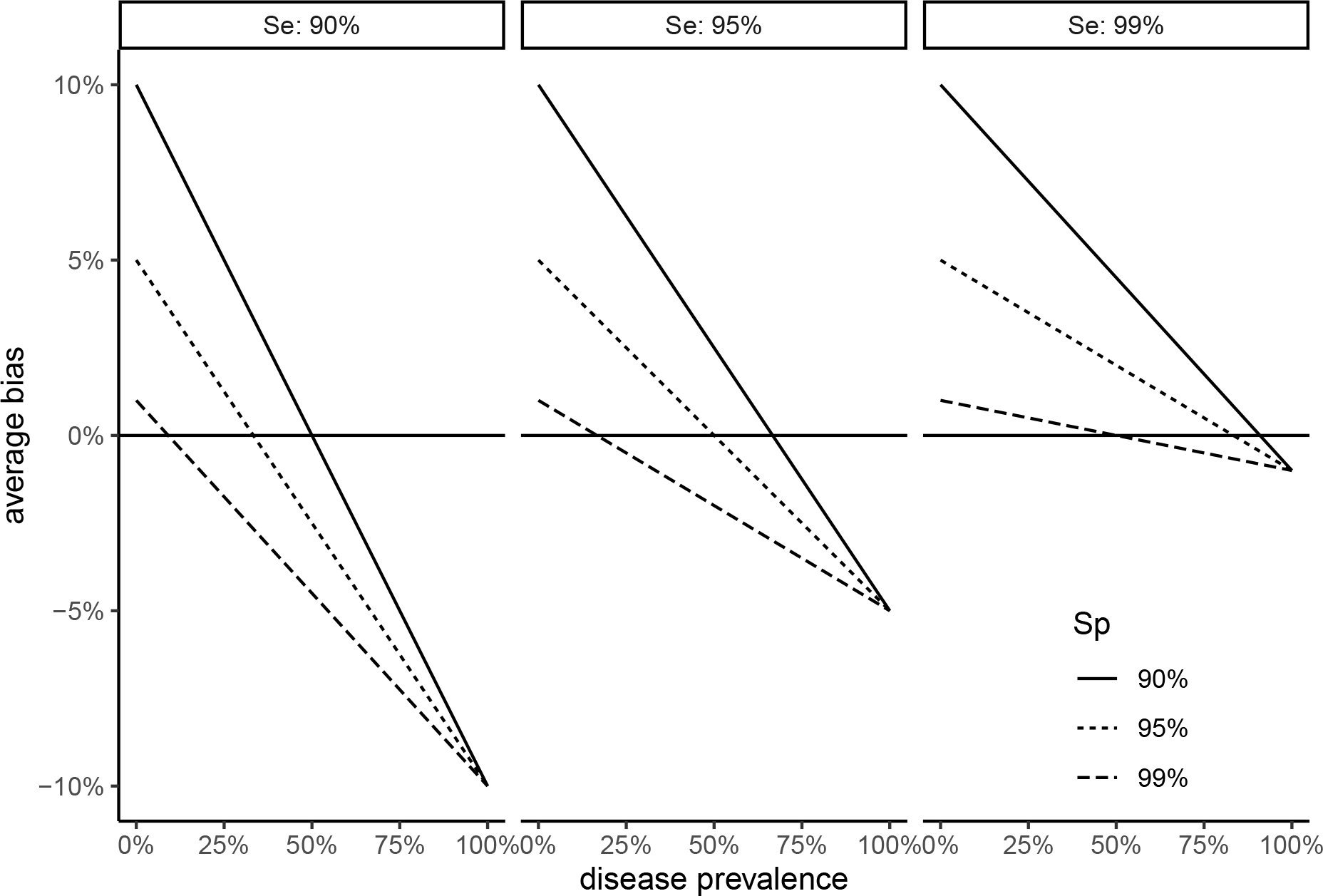
Estimated bias in prevalence estimate for selected combinations of sensitivity, specificity and true disease prevalence

**Figure 2:**
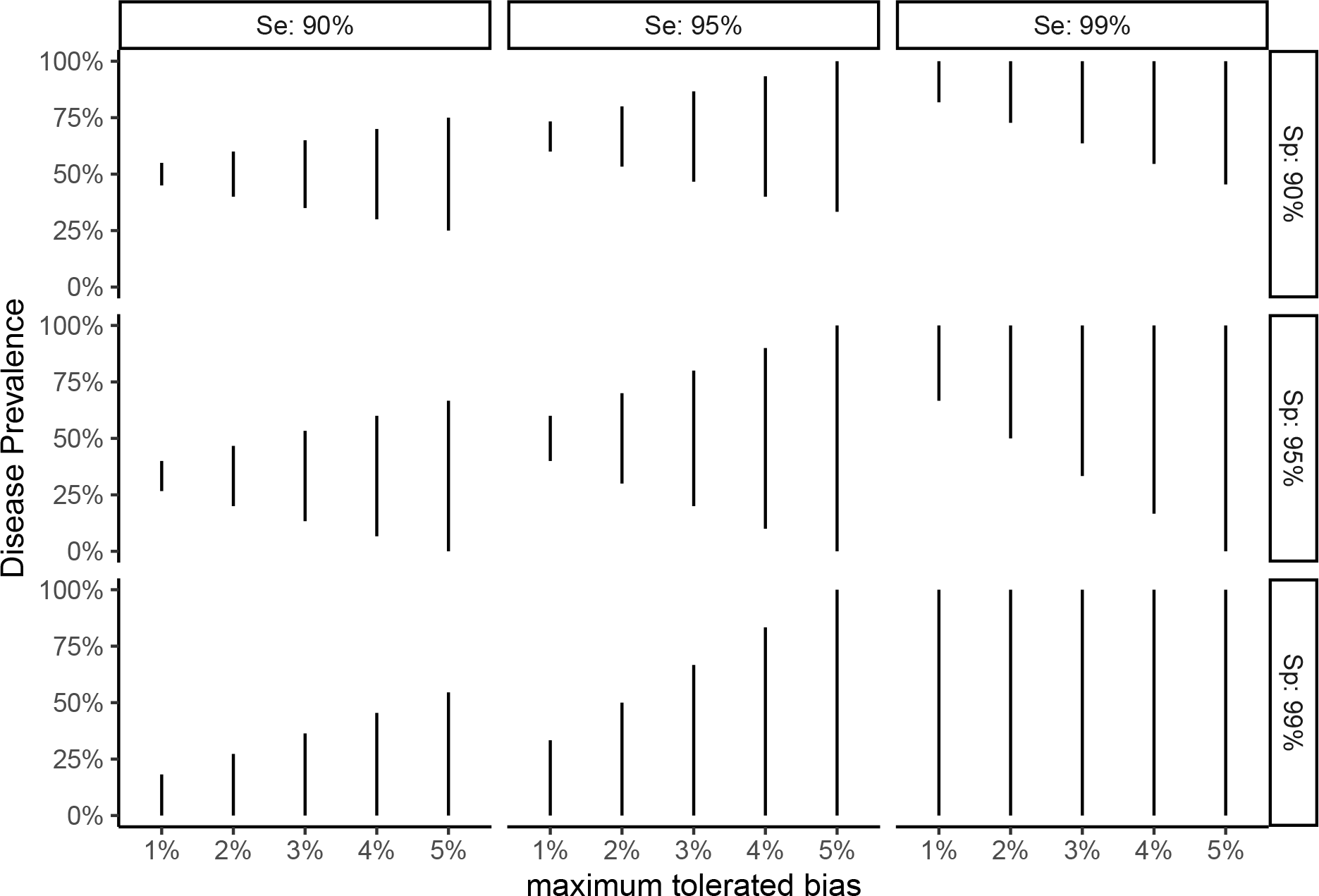
Range of true disease prevalence where the rate of positive tests is a close approximation of disease prevalence, to within maximum absolute tolerated bias

Using the bounds of prevalence as derived above, we explored where the maximum tolerated bias is limited to 1%, 2.5% and 5% (Figure 2). When *Se* and *Sp* are each 90%, bias is within a tolerance of 1% only very close to 50% disease prevalence, within 2.5% tolerance in the range of 38% - 62% disease prevalence and to within 5% tolerance as long as disease prevalence is between 25% and 75%. When the desired tolerance is 1%, the range of disease prevalence where a naive approach will yield unbiased results is fairly narrow in all cases, unless *Se* and *Sp* are each at least 99%. Outside of these ranges, using the proportion of positive test results to estimate seroprevalence will be too biased, and more sophisticated analysis methods should be used.

As an example of this, take the Ciao Corona study (19), a school-based longitudinal study of seroprevalence in Swiss school children with 5 rounds of SARS-CoV-2 antibody testing between June 2020 and June 2022. The antibody test used has a sensitivity of 94% in children, and a specificity of 99.2%. In June 2020, 98 / 2473 (4.0%) of subjects showed as seropositive, compared to 154 / 2500 (6.2%) in October 2021, 17.3% (426 / 2453) in March 2021, 48.5% (910 / 1876) in November 2021, and 94.5% (2008 / 2125) in June 2022. Given the diagnostic test characteristics, absolute bias can be expected to be less than 1% in the range of 0% - 26.5% disease prevalence, and less than 2% for disease prevalence of up to 41.2%. These results imply that reported seroprevalence estimates based on a naive logistic approach are likely relatively unbiased for the first 3 rounds of Ciao Corona antibody testing (0.5%, 0.4% and -0.4% respectively), but that after that any seroprevalence estimates that do not adjust for test characteristics are likely quite biased (-2.4% and -5.6%). In order to adjust for covariates and survey sampling weights, we corrected the seroprevalence estimates using a Bayesian hierarchical model approach in all rounds of testing.

## Discussion

We have demonstrated that average bias in prevalence estimates can be higher than desired, as high as 10%, when using a naive approach of calculation based on the proportion of positive test results, even if sensitivity and specificity are 90% or higher. Further, we have derived a range of disease prevalence values for which the naive approach gives reasonably unbiased prevalence estimates. A systematic review indicates that many public health researchers are not aware of methods for reducing this potential bias, and do not correct for this in their own studies of prevalence. Nor do peer reviewers and editors seem to notice this widespread problem.

Taken together, the results emphasize the necessity in public health research to not simply report raw proportions of positive tests, even if those are adjusted for demographic characteristics using logistic regression. Since disease prevalence is of course not known precisely prior to study conduct, the most straightforward approach is then to plan statistical methods so that sensitivity and specificity are accounted for. Even if other sources of bias (e.g. sampling bias, or sampling variation) are accounted for, the results of seroprevalence studies will continue to be biased if analyses do not also account for test sensitivity and specificity. Care should also be taken in reading publications reporting (sero)prevalence estimates to ensure that suitable statistical methods have been used.

These results are based on the definitions of sensitivity and specificity only and require no complicated derivations. we have not adjusted for demographic characteristics, such as age and gender, or used weighting to approximate the target population, as is typical in surveys of disease prevalence. However, such adjustment cannot alleviate any general concerns of bias as presented here. The bias demonstrated here is also an average bias, and observed bias may vary more or less depending on the size of the sample. The results do not account for other possible issues with a diagnostic test (20–22), that can often not be corrected with statistical methods (e.g. when the validation sample, on which the sensitivity and specificity estimates are based, is not similar to the population of interest). Average bias is given, which does not account for sampling bias or variation, as has been described elsewhere (9).

The question remains as to how best to account for diagnostic test sensitivity and specificity when estimating disease prevalence. A nice outline of some appropriate methods along with implementation in R (23) code is given by (13,24,25). To calculate corrected confidence intervals for prevalence in studies where covariates do not need to be adjusted for, and no survey weights are needed, the R package bootComb (26) and website “epitools” (https://epitools.ausvet.com.au/trueprevalence) are available, while Bayesian methods are available in prevalence (27). Using the Rogan-Gladen correction with bootstrap confidence intervals, or the Bayesian correction in the prevalence package are appropriate when there is no need to adjust for any other factors. Adjusting for covariates, adjusting for sampling bias or variation, or application of post-stratification weights (among other issues) may unfortunately need to be done without the use of such prepackaged code, e.g. as described by (28). Collaboration with experienced statisticians is invaluable in ensuring that correct analysis techniques are used so that unbiased prevalence estimates can be reported.

The majority of publications, even if high impact journals, reporting seroprevalence estimates in the literature do not account for sensitivity and specificity of the diagnostic test. Bias introduced by reporting the proportion of positive tests rather than prevalence can be easily as high as 10%, or more if sensitivity or specificity are less than 90%. Public health researchers performing prevalence studies should consult experienced statisticians when analyzing such data, and be sure to account for test performance. However, researchers reviewing published prevalence studies also need to be aware of this issue. The results here will assist reviewers in determining the the magnitude of bias that can be expected, so that publications in the epidemiology literature can be interpreted properly.

## Data Availability

All data produced in the present study are available upon reasonable request to the authors.

## Funding statement

The Ciao Corona study, used in our example, is part of Corona Immunitas research network, coordinated by the Swiss School of Public Health (SSPH+), and funded by fundraising of SSPH+ that includes funds of the Swiss Federal Office of Public Health and private funders (ethical guidelines for funding stated by SSPH+ will be respected), by funds of the Cantons of Switzerland (Vaud, Zurich, and Basel) and by institutional funds of the Universities. Additional funding, specific to this study is available from the University of Zurich Foundation. The EBPI at the University of Zurich provided funding for the systematic review. The funders had no involvement in the systematic review, writing of this report, or decision to submit the paper for publication.

## Competing interests

The authors declare no competing interests.

## Contributions

SRH initiated the analysis, developed the methodology, planned and conducted the systematic review, performed the statistical analysis, and wrote the manuscript. DK, in parallel with SRH, reviewed publications for the systematic review, extracted relevant data, and reviewed the manuscript. No others meeting criteria for authorship have been omitted.

## Acknowledgements

The authors thank Julia Braun, Thomas Radtke, and Milo Puhan for their critical comments.

## Notes

### Competing Interest Statement

The authors have declared no competing interest.

### Author Declarations

The study was approved by the Ethics Committee of the Canton of Zurich, Switzerland (2020-01336).

### Summary of Updates

References to Figures 1 and 2 were corrected, and references to Figure 3 were removed.

