## Supplementary Material for "Potential for bias in (sero)prevalence estimates when not accounting for test sensitivity and specificity"

### List of Tables

S1 Summary of inclusion / exclusion decisions for the systematic review. . . . . 1
S2 Key outcomes of systematic review. The sensitivity analysis included 316 publications meeting all inclusion criteria, plus any research letters meeting all other inclusion criteria. . . . . . . . . . . . . . . . . . . . . . . . . . . 2

Table S1: Summary of inclusion / exclusion decisions for the systematic review.

| **Characteristic** | **N = 640** |
|---|---|
| inclusion | |
| Duplicate | 4 (0.6%) |
| Excluded: conflict of interest | 5 (0.8%) |
| Excluded: foreign language | 7 (1.1%) |
| Excluded: not Covid-19 | 2 (0.3%) |
| Excluded: not in humans | 9 (1.4%) |
| Excluded: secondary research | 22 (3.4%) |
| Excluded: did not assess seroprevalence | 233 (36%) |
| Excluded: risk factors or subgroups | 41 (6.4%) |
| Excluded: no full text | 3 (0.5%) |
| Sensitivity analysis: research letter | 23 (3.6%) |
| Included | 291 (45%) |

Table S2: Key outcomes of systematic review. The sensitivity analysis included 316 publications meeting all inclusion criteria, plus any research letters meeting all other inclusion criteria.

| **Characteristic** | **Overall**<br>N = 314 (100%) | **uncorrected**<br>N = 244 (78%) | **corrected**<br>N = 70 (22%) |
|---|---|---|---|
| Statistical method | | | |
| Rogan-Gladen | 40 (13%) | 0 (0%) | 40 (57%) |
| Bayesian | 22 (7.0%) | 0 (0%) | 22 (31%) |
| unspecified method | 8 (2.5%) | 0 (0%) | 8 (11%) |
| unclear | 2 (0.6%) | 2 (0.8%) | 0 (0%) |
| uncorrected | 242 (77%) | 242 (99%) | 0 (0%) |
| Reported test Se and Sp? | | | |
| did not report | 114 (36%) | 109 (45%) | 5 (7.1%) |
| partially reported | 11 (3.5%) | 11 (4.5%) | 0 (0%) |
| reported | 189 (60%) | 124 (51%) | 65 (93%) |
| Se | 95.1 (60.2 - 100.0) | 96.8 (60.2 - 100.0) | 92.7 (66.9 - 100.0) |
| Sp | 99.6 (82.4 - 100.0) | 99.5 (86.5 - 100.0) | 99.8 (82.4 - 100.0) |
| expected bias | 0.0 (-12.2 - 9.1) | 0.0 (-12.2 - 7.1) | -0.3 (-12.2 - 9.1) |
| expected bias (category) | | | |
| [-15,-10) | 5 (2.6%) | 3 (2.4%) | 2 (3.1%) |
| [-10,-5) | 8 (4.2%) | 4 (3.2%) | 4 (6.2%) |
| [-5,-1) | 29 (15%) | 18 (15%) | 11 (17%) |
| [-1,1) | 116 (61%) | 75 (60%) | 41 (63%) |
| [1,5) | 28 (15%) | 22 (18%) | 6 (9.2%) |
| [5,10] | 3 (1.6%) | 2 (1.6%) | 1 (1.5%) |

[1] n (%); Median (Range)
